# Predicting critical state after COVID-19 diagnosis: Model development using a large US electronic health record dataset

**DOI:** 10.1101/2020.07.24.20155192

**Authors:** Mike D. Rinderknecht, Yannick Klopfenstein

**Affiliations:** IBM Switzerland Ltd, Zurich, Switzerland

**Keywords:** AI, artificial intelligence, clinical decision support, coronavirus, IBM Explorys, machine learning, prognosis, real-world evidence, risk stratification, RWE, SARS-CoV-2, triage

## Abstract

As the COVID-19 pandemic is challenging healthcare systems worldwide, early identification of patients with a high risk of complication is crucial. We present a prognostic model predicting critical state within 28 days following COVID-19 diagnosis trained on data from US electronic health records (IBM Explorys), including demographics, comorbidities, symptoms, insurance types, and hospitalization. Out of 15816 COVID-19 patients, 2054 went into critical state or deceased. Random, stratified train-test splits were repeated 100 times and lead to a ROC AUC of 0.872 [0.868, 0.877] and a precision-recall AUC of 0.500 [0.488, 0.509] (median and interquartile range). The model was well-calibrated, showing minor tendency to over-forecast probabilities above 0.5. The interpretability analysis confirmed evidence on major risk factors (e.g., older age, higher BMI, male gender, diabetes, and cardiovascular disease) in an efficient way compared to clinical studies, demonstrating the model validity. Such personalized predictions could enable fine-graded risk stratification for optimized care management.

## 1 Introduction

The coronavirus disease (COVID-19), caused by the severe acute respiratory syndrome coronavirus 2 (SARS-CoV-2) ^[1]^, has started to spread since December 2019 from the province Hubei of the People’s Republic of China to 188 countries, becoming a global pandemic ^[2]^. Despite having a lower case fatality rate than SARS in 2003 and MERS in 2012 ^[3]^, the overall number of 24 007 049 cases and 821933 deaths from COVID-19 ^[2]^ (status August 26, 2020) far outweigh the other two epidemics. These high numbers have forced governments to respond with severe containment strategies to delay the spread of COVID-19 in order to avoid a global health crisis and collapse of the healthcare systems ^[4,5]^. Several countries have been facing shortages of intensive care beds or medical equipment such as ventilators ^[6]^. Given these circumstances, appropriate prognostic tools for identifying high-risk populations and helping triage are essential for informed protection policies by policymakers and optimal resource allocation to ensure best possible and early care for the patients.

Today’s availability of data enables the development of different solutions using machine learning to address these needs, as described in recent reviews ^[7,8]^. One type of proposed solutions is prognostic prediction modeling, which consists in predicting patient outcomes such as hospitalization or exacerbation to a critical state, using longitudinal data from medical healthcare records of COVID-19 patients ^[9–19]^ or proxy datasets based on other upper respiratory infections ^[20]^. To this date, most studies include data exclusively from one or few hospitals and therefore relatively small sample sizes of COVID-19 patients (i.e., below 1000 patients), with the exception of the retrospective studies in New York City with 4103 ^[16]^ or with a total of 3055 patients ^[17]^.

This is where combined electronic health records (EHRs) across a large network of hospitals and care providers become valuable to generate real-world evidence (RWE). Machine learning models based on such datasets can benefit from increased amount of data and improved robustness and generalizability, as data comes from various sources (e.g., different hospitals), and may thus cover wider ranges of demographics and diverse healthcare practices or systems. Having such data available, can facilitate and accelerate insight generation, as such an approach for retrospective data analyses is more cost effective and requires less effort compared to setting up and running large scale clinical studies.

The aim of this work was to create a prognostic prediction model for critical state after COVID-19 diagnosis based on a retrospective analysis of a large set of de-identified EHRs of patients across the US using the IBM® Explorys® database (IBM, Armonk, NY). Such a predictive model allows identifying patients at risk based on predictive factors to support risk stratification and enable early triage. The present work based on EHR data is reported according to the RECORD and STROBE statements ^[21]^ and reporting of model development followed TRIPOD statement guidelines ^[22]^.

## 2 Results

### 2.1 Cohort, descriptive statistics, and concurvity

The total number of identified patients diagnosed with COVID-19 based on International Classification of Diseases (ICD) codes and entries for positive results of SARS-CoV-2 test based on Logical Observation Identifiers Names and Codes (LOINC) are reported in **Figure 1**. In addition, the number of patients with age and gender information (referred to as the cohort), the number of patients labeled as not entering critical state and labeled as entering critical state as well as the sizes of the partitions for training and testing are also reported. Among patients labeled as critical state, a total of 545 patients were flagged as deceased in the Explorys database. This corresponds to 3.4% of the entire cohort.

**Figure 1.**
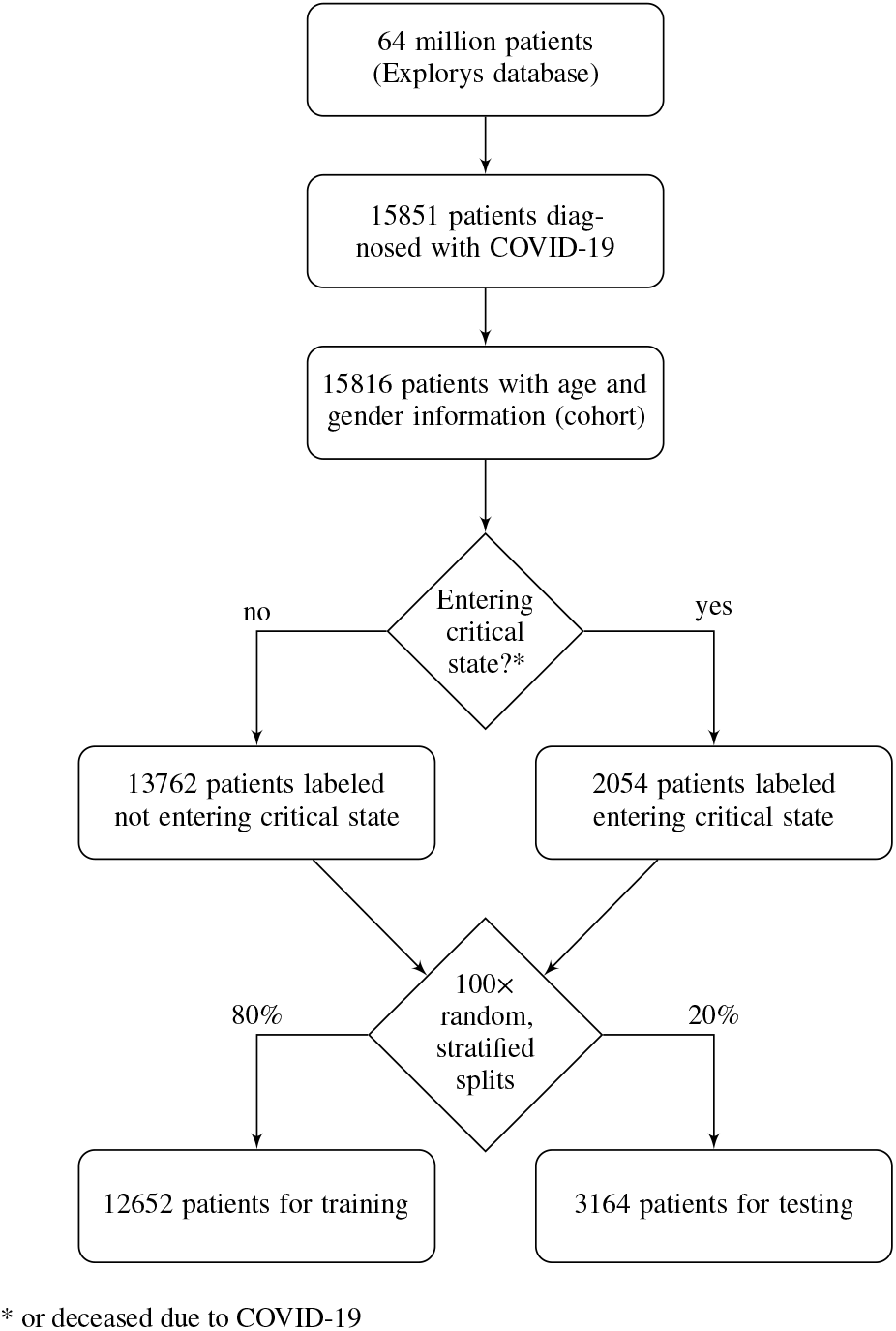
Diagram of number of subjects. Cohort selection and number of patients not entering versus entering critical state based on the definitions outlined in the according sections. To train and evaluate the model, the dataset was split using stratification of the prediction target. This procedure was repeated 100 times based on random seeds to get a distribution of model performance.

Descriptive statistics after zero-imputation of the binary features and before feature reduction are reported in **Table 1**. No features were removed due to a too high proportion of missing data. Rank correlations across features is shown in the heatmap in **Figure 2**. The following feature combination showed a strong rank correlation: {Race (African American), Race (Caucasian)}. The following feature was removed to avoid feature collinearity: Race (Caucasian). The removed feature will therefore contribute to the baseline risk probability.

**Figure 2.**
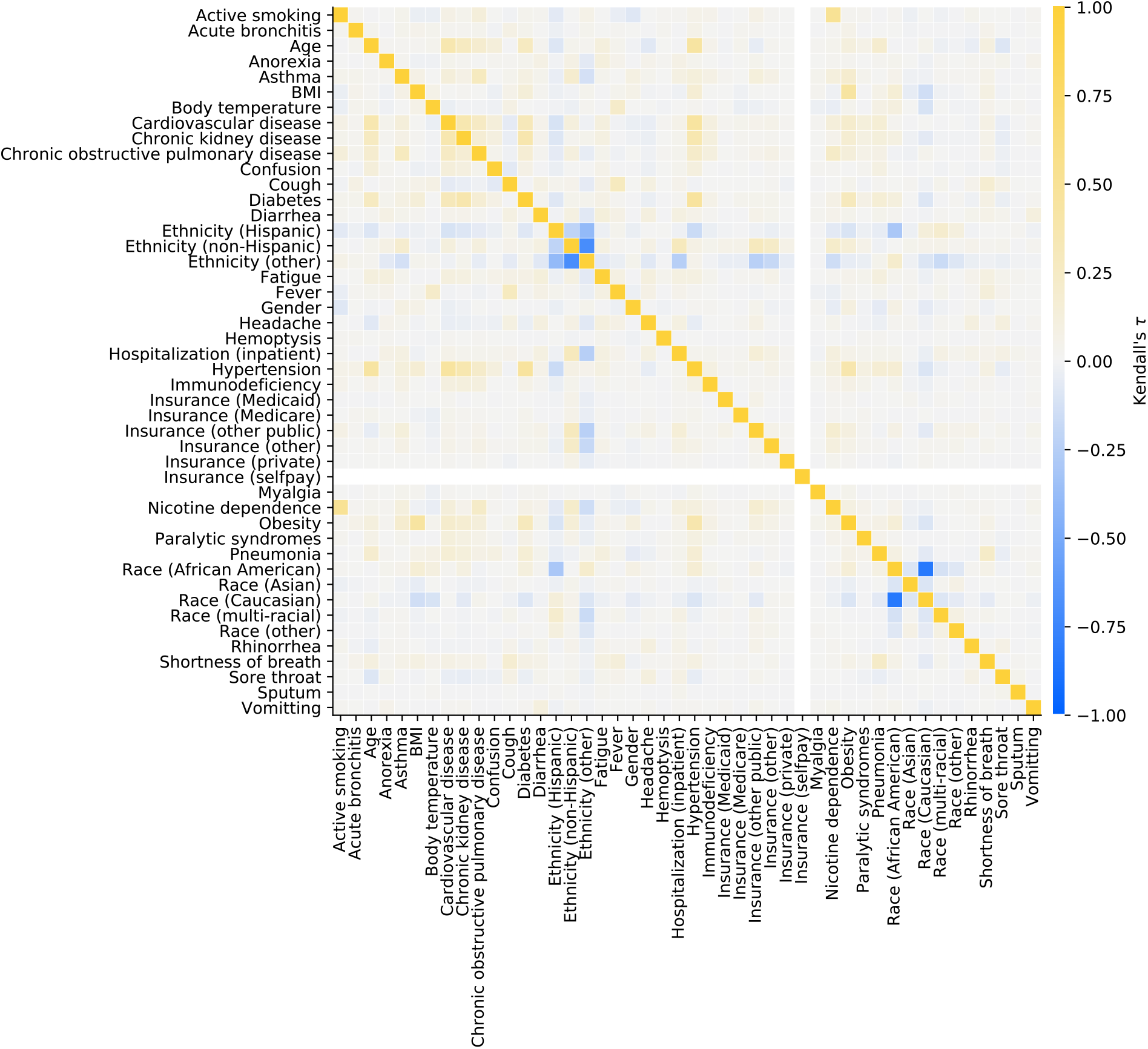
Feature concurvity. Kendall’s **t** was used to evaluate correlation between each feature combination. If all patients of the entire cohort have the same feature value, no concurvity can be calculated (represented in white, e.g., for Insurance (selfpay)).Calibration curve

**Table 1.**
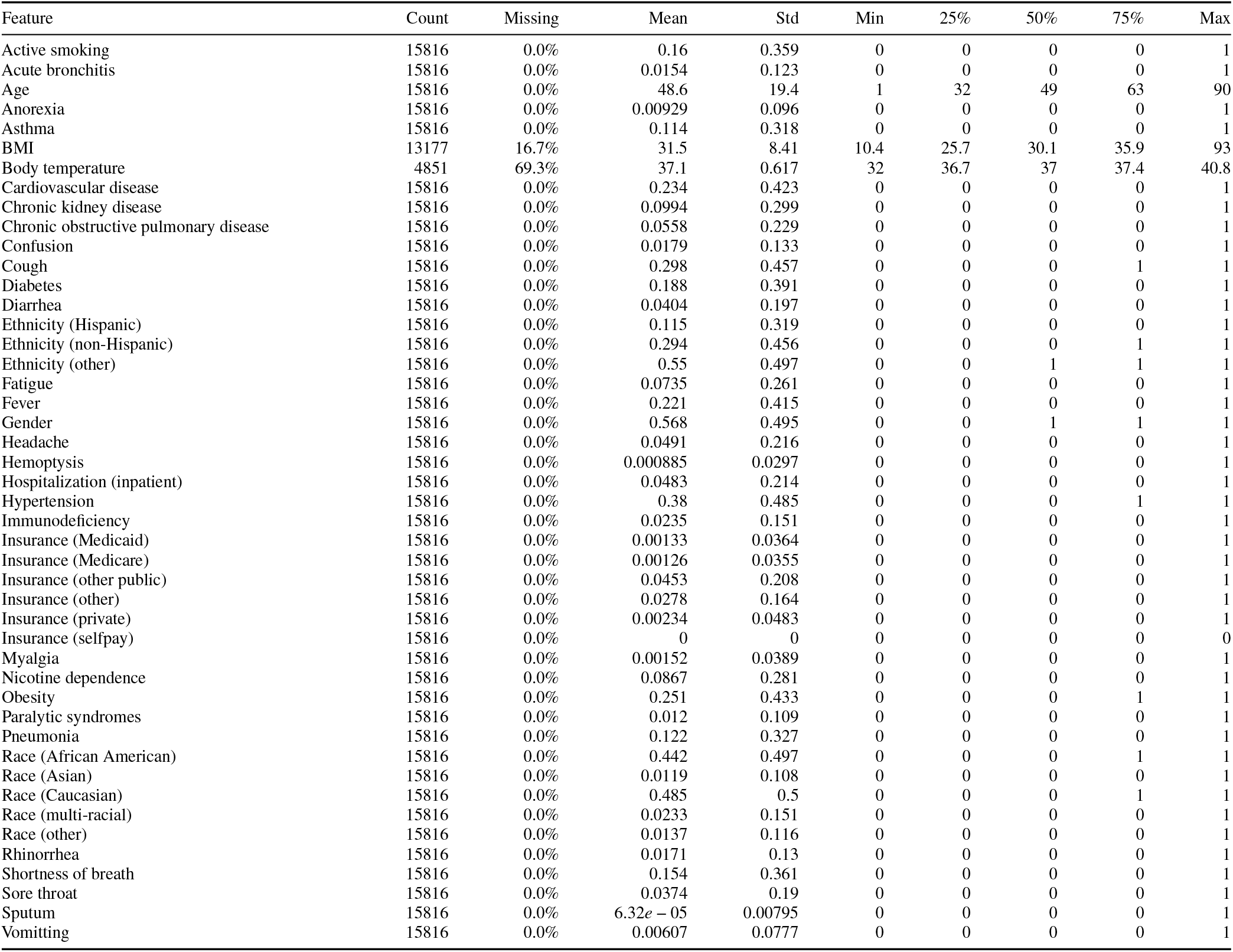
Descriptive statistics of the features. The descriptive statistics are based on the full dataset after zero-imputation of the binary features but before feature reduction. The percentages 25%, 50%, and 75% refer to the first (Q1), second (median), and third quartiles (Q3). Note that for binary features the Mean column represents the proportion of positive entries. Note that as part of Explorys’ de-identification process the feature Age has a ceiling effect at 90 years, and the age of all patients born in the last 365 days is reported as zero. For gender, 1 corresponds to female.

| Feature | Count | Missing | Mean | Std | Min | 25% | 50% | 75% | Max |
| --- | --- | --- | --- | --- | --- | --- | --- | --- | --- |
| Active smoking | 15816 | 0.0% | 0.16 | 0.359 | 0 | 0 | 0 | 0 | 1 |
| Acute bronchitis | 15816 | 0.0% | 0.0154 | 0.123 | 0 | 0 | 0 | 0 | 1 |
| Age | 15816 | 0.0% | 48.6 | 19.4 | 1 | 32 | 49 | 63 | 90 |
| Anorexia | 15816 | 0.0% | 0.00929 | 0.096 | 0 | 0 | 0 | 0 | 1 |
| Asthma | 15816 | 0.0% | 0.114 | 0.318 | 0 | 0 | 0 | 0 | 1 |
| BMI | 13177 | 16.7% | 31.5 | 8.41 | 10.4 | 25.7 | 30.1 | 35.9 | 93 |
| Body temperature | 4851 | 69.3% | 37.1 | 0.617 | 32 | 36.7 | 37 | 37.4 | 40.8 |
| Cardiovascular disease | 15816 | 0.0% | 0.234 | 0.423 | 0 | 0 | 0 | 0 | 1 |
| Chronic kidney disease | 15816 | 0.0% | 0.0994 | 0.299 | 0 | 0 | 0 | 0 | 1 |
| Chronic obstructive pulmonary disease | 15816 | 0.0% | 0.0558 | 0.229 | 0 | 0 | 0 | 0 | 1 |
| Confusion | 15816 | 0.0% | 0.0179 | 0.133 | 0 | 0 | 0 | 0 | 1 |
| Cough | 15816 | 0.0% | 0.298 | 0.457 | 0 | 0 | 0 | 1 | 1 |
| Diabetes | 15816 | 0.0% | 0.188 | 0.391 | 0 | 0 | 0 | 0 | 1 |
| Diarrhea | 15816 | 0.0% | 0.0404 | 0.197 | 0 | 0 | 0 | 0 | 1 |
| Ethnicity (Hispanic) | 15816 | 0.0% | 0.115 | 0.319 | 0 | 0 | 0 | 0 | 1 |
| Ethnicity (non-Hispanic) | 15816 | 0.0% | 0.294 | 0.456 | 0 | 0 | 0 | 1 | 1 |
| Ethnicity (other) | 15816 | 0.0% | 0.55 | 0.497 | 0 | 0 | 1 | 1 | 1 |
| Fatigue | 15816 | 0.0% | 0.0735 | 0.261 | 0 | 0 | 0 | 0 | 1 |
| Fever | 15816 | 0.0% | 0.221 | 0.415 | 0 | 0 | 0 | 0 | 1 |
| Gender | 15816 | 0.0% | 0.568 | 0.495 | 0 | 0 | 1 | 1 | 1 |
| Headache | 15816 | 0.0% | 0.0491 | 0.216 | 0 | 0 | 0 | 0 | 1 |
| Hemoptysis | 15816 | 0.0% | 0.000885 | 0.0297 | 0 | 0 | 0 | 0 | 1 |
| Hospitalization (inpatient) | 15816 | 0.0% | 0.0483 | 0.214 | 0 | 0 | 0 | 0 | 1 |
| Hypertension | 15816 | 0.0% | 0.38 | 0.485 | 0 | 0 | 0 | 1 | 1 |
| Immunodeficiency | 15816 | 0.0% | 0.0235 | 0.151 | 0 | 0 | 0 | 0 | 1 |
| Insurance (Medicaid) | 15816 | 0.0% | 0.00133 | 0.0364 | 0 | 0 | 0 | 0 | 1 |
| Insurance (Medicare) | 15816 | 0.0% | 0.00126 | 0.0355 | 0 | 0 | 0 | 0 | 1 |
| Insurance (other public) | 15816 | 0.0% | 0.0453 | 0.208 | 0 | 0 | 0 | 0 | 1 |
| Insurance (other) | 15816 | 0.0% | 0.0278 | 0.164 | 0 | 0 | 0 | 0 | 1 |
| Insurance (private) | 15816 | 0.0% | 0.00234 | 0.0483 | 0 | 0 | 0 | 0 | 1 |
| Insurance (selfpay) | 15816 | 0.0% | 0 | 0 | 0 | 0 | 0 | 0 | 0 |
| Myalgia | 15816 | 0.0% | 0.00152 | 0.0389 | 0 | 0 | 0 | 0 | 1 |
| Nicotine dependence | 15816 | 0.0% | 0.0867 | 0.281 | 0 | 0 | 0 | 0 | 1 |
| Obesity | 15816 | 0.0% | 0.251 | 0.433 | 0 | 0 | 0 | 1 | 1 |
| Paralytic syndromes | 15816 | 0.0% | 0.012 | 0.109 | 0 | 0 | 0 | 0 | 1 |
| Pneumonia | 15816 | 0.0% | 0.122 | 0.327 | 0 | 0 | 0 | 0 | 1 |
| Race (African American) | 15816 | 0.0% | 0.442 | 0.497 | 0 | 0 | 0 | 1 | 1 |
| Race (Asian) | 15816 | 0.0% | 0.0119 | 0.108 | 0 | 0 | 0 | 0 | 1 |
| Race (Caucasian) | 15816 | 0.0% | 0.485 | 0.5 | 0 | 0 | 0 | 1 | 1 |
| Race (multi-racial) | 15816 | 0.0% | 0.0233 | 0.151 | 0 | 0 | 0 | 0 | 1 |
| Race (other) | 15816 | 0.0% | 0.0137 | 0.116 | 0 | 0 | 0 | 0 | 1 |
| Rhinorrhea | 15816 | 0.0% | 0.0171 | 0.13 | 0 | 0 | 0 | 0 | 1 |
| Shortness of breath | 15816 | 0.0% | 0.154 | 0.361 | 0 | 0 | 0 | 0 | 1 |
| Sore throat | 15816 | 0.0% | 0.0374 | 0.19 | 0 | 0 | 0 | 0 | 1 |
| Sputum | 15816 | 0.0% | 6.32e-05 | 0.00795 | 0 | 0 | 0 | 0 | 1 |
| Vomitting | 15816 | 0.0% | 0.00607 | 0.0777 | 0 | 0 | 0 | 0 | 1 |

**Table 2.** ICD-10 codes for the prediction target. Patients with first diagnosis of any of the listed ICD-10 codes within the specified time window were labeled as entering critical state.

| ICD-10 code | Description |
| --- | --- |
| A41.89 | Other specified sepsis |
| A41.9 | Sepsis, unspecified organism |
| R65.2 | Severe sepsis |
| R65.20 | Severe sepsis without septic shock |
| R65.21 | Severe sepsis with septic shock |
| J80 | Acute respiratory distress syndrome (ARDS) |
| J96 | Respiratory failure, not elsewhere classified |
| J96.0 | Acute respiratory failure |
| J96.00 | Acute respiratory failure, unspecified whether with hypoxia or hypercapnia |
| J96.01 | Acute respiratory failure with hypoxia |
| J96.02 | Acute respiratory failure with hypercapnia |
| J96.9 | Respiratory failure, unspecified |
| J96.90 | Respiratory failure, unspecified, unspecified whether with hypoxia or hypercapnia |
| J96.91 | Respiratory failure, unspecified with hypoxia |
| J96.92 | Respiratory failure, unspecified with hypercapnia |

### 2.2 Performance

The performance and calibration of the model was evaluated on the 3164 patients of the test set for each train-test split seed. The area under the curve (AUC) of the receiver operating characteristic (ROC) and under the precision-recall (PR) curve across the 100 different seeds were 0.872 [0.868, 0.877] and 0.500 [0.488, 0.509], respectively (median and interquartile range). **Figure 3** shows their distributions, together with the ROC curve and the precision recall curve (Left and Middle), as well as the calibration of the model (Right). The confusion matrix for the identified optimal classification threshold (0.127 [0.118, 0.139]) is shown in **Figure 4**. The sensitivity of the model for the optimal threshold was 0.841 [0.819, 0.856] and the specificity 0.762 [0.740, 0.781].

**Figure 3.**
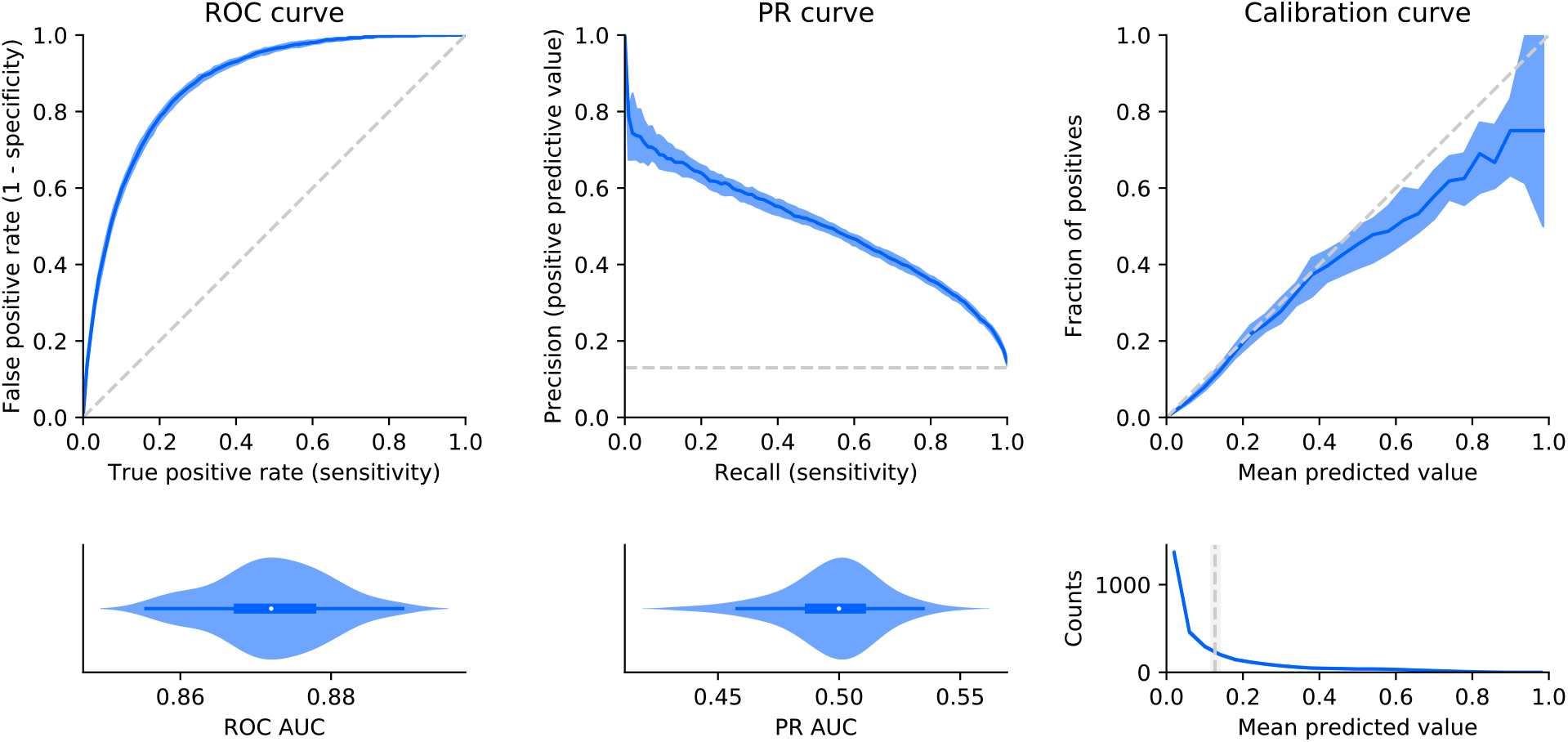
Model performance and calibration. Left: Receiver operating characteristic (ROC) curve and corresponding normalized violin plot of the distribution of the ROC area under the curve (AUC). The top plot shows median and interquartile range (IQR) of the performance (blue) and the chance level (no predictive value) as a reference (dashed gray line). Middle: Same representation for the precision recall (PR) curve and corresponding distribution of the PR AUC. Right: Median and IQR (blue) of the fraction of actual positives (labeled as critical state) for the binned mean predicted values (i.e., probabilities) (top). The reference diagonal represents perfect calibration (dashed gray line). The bottom plot shows median and IQR (blue) for the counts within each bin of mean predicted value. The vertical gray line shows the median and IQR for the optimal decision threshold based on the Youden’s J statistic.

**Figure 4.**
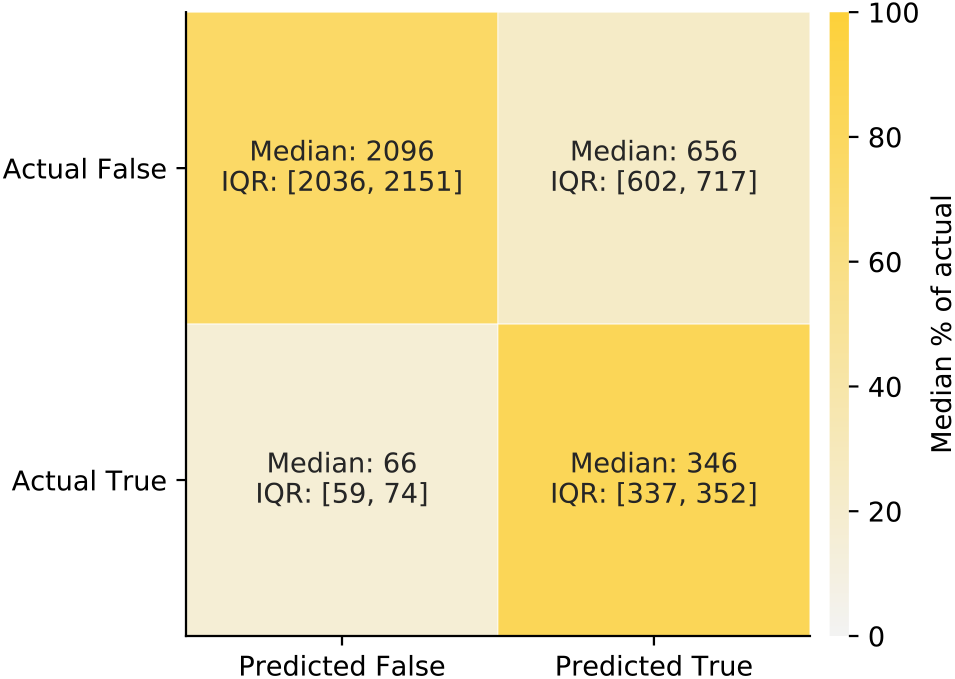
Confusion matrix. Confusion matrix for the predictions of the test set based on the optimal decision threshold. True refers to entering critical state, and False refers to not entering critical state. The shades of the confusion matrix correspond to the median percentage of the actual labels (i.e., shade of the top left cell and the bottom right cell represent the median specificity and the median sensitivity, respectively).

### 2.3 Model interpretability

**Figure 5** shows the results of the model interpretability analysis based on Tree SHAP ^[23]^. Older age and pneumonia are by far the principal predictors for critical state. The main features contributing to a higher probability of critical state in case of high feature values or presence are (in decreasing order of global feature importance): older age, pneumonia, higher BMI, diabetes, male gender, shortness of breath, and cardiovascular disease. Note that in **Figure 5** for binary features “max” feature values correspond to 1 (e.g., presence of the feature). In the case of gender, 1 corresponds to female (see **Table 3**). **Figure 6** illustrates the composition of a single example prediction.

**Figure 5.**
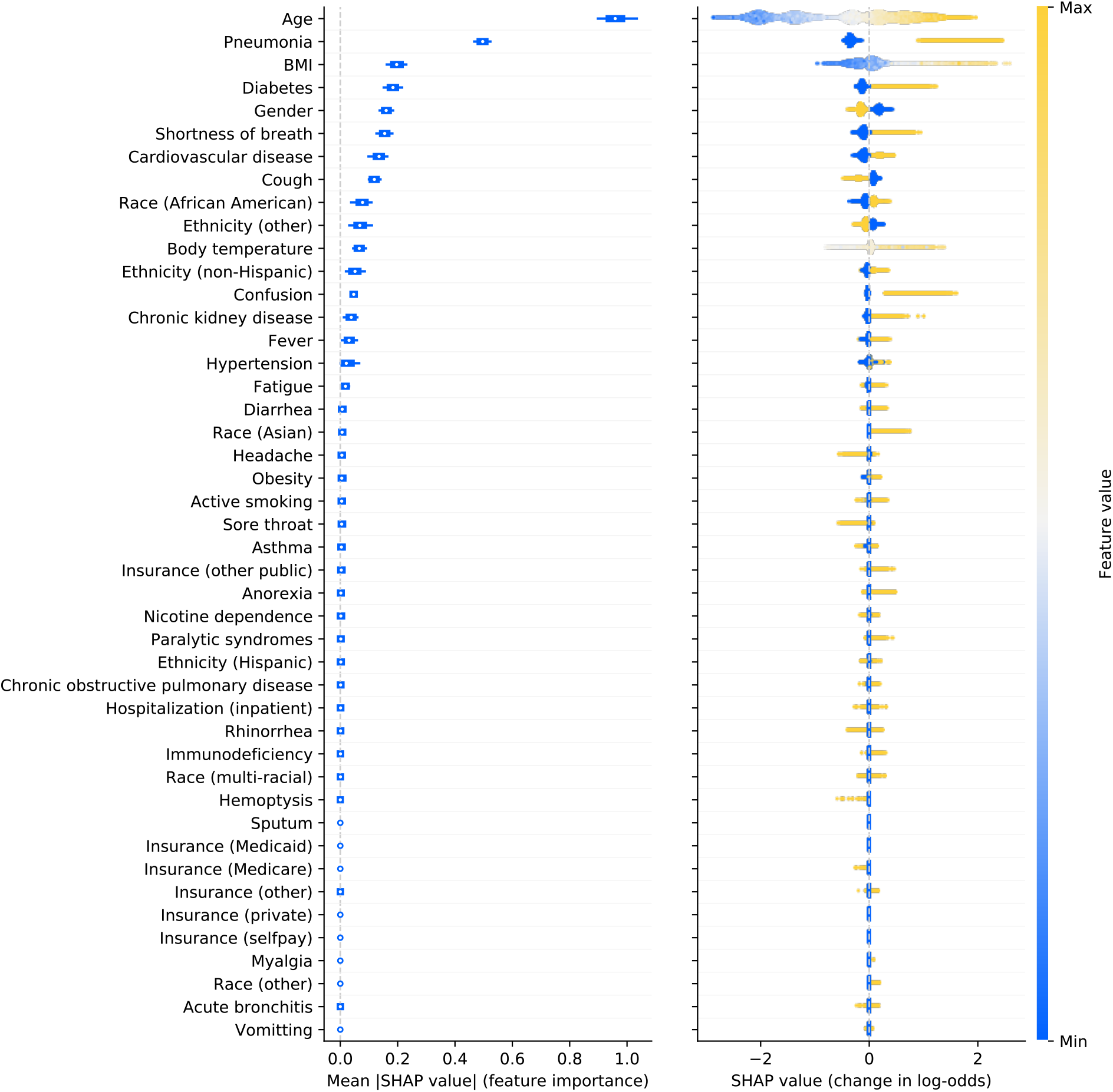
Model interpretability. Left: Box plots (across different seeds) of the average absolute impact of features on the model output magnitude (in log-odds) ordered by decreasing feature importance. Boxplots indicate median (circle), interquartile range (box), Tukey’s original definition for the whiskers (1.5 times the interquartile range). Right: Illustration of the relation between feature values and impact (in terms of magnitude and direction) on prediction output (all seeds pooled). Each dot represents an individual patient in the test set. The color of each point corresponds to the normalized feature value (min-max normalization on test set). As an example for continuous features, older patients tend to have a higher SHAP value). For binary features, the maximum feature value 1 corresponds to presence of the feature, and 0 to absence of the feature. For gender, 1 corresponds to female and 0 to male.

**Figure 6.**
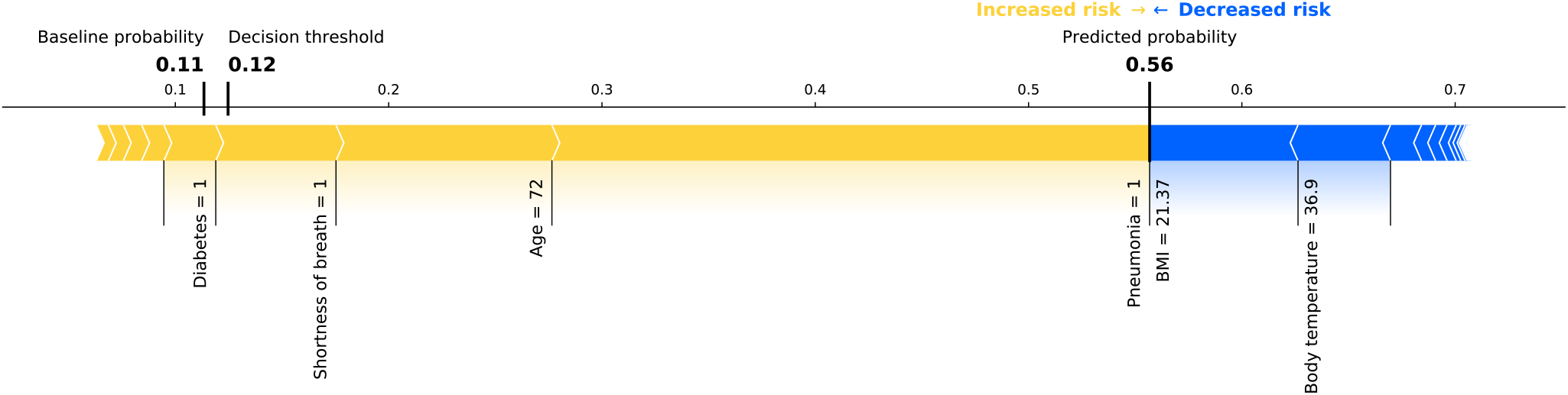
Example prediction. Composition of a prediction (in probability space) for an example patient going into critical state, based on the model of a random seed. The yellow arrows represent the contributions of major risk factors (e.g., older age or diabetes), and the blue arrows represent the contribution of factors decreasing the probability of entering critical state (e.g., healthy BMI and normal body temperature). Note that the baseline probability and the decision threshold are relatively low and close to the actual ratio of patients entering critical state in our cohort.

**Table 3.** Feature definitions. Feature names, units and details (e.g., ICD and LOINC codes) grouped by extraction time window specifications.

| Extraction time window | Feature | Units | Details |
| --- | --- | --- | --- |
| Special features | Age | Years | Computed at diagnosis date, based on birth year entry |
|  | Gender | NA (0: male, 1: female) | No time window restrictions |
|  | Ethnicity (Hispanic) | NA (binary) | No time window restrictions |
|  | Ethnicity (non-Hispanic) | NA (binary) | No time window restrictions |
|  | Ethnicity (Other) | NA (binary) | No time window restrictions |
|  | Insurance (Medicaid) | NA (binary) | No time window restrictions |
|  | Insurance (Medicare) | NA (binary) | No time window restrictions |
|  | Insurance (Other) | NA (binary) | No time window restrictions |
|  | Insurance (Other public) | NA (binary) | No time window restrictions |
|  | Insurance (Private) | NA (binary) | No time window restrictions |
|  | Insurance (Selfpay) | NA (binary) | No time window restrictions |
|  | Race (African American) | NA (binary) | No time window restrictions |
|  | Race (Asian) | NA (binary) | No time window restrictions |
|  | Race (Caucasian) | NA (binary) | No time window restrictions |
|  | Race (Multi-racial) | NA (binary) | No time window restrictions |
|  | Race (Other) | NA (binary) | No time window restrictions |
| Acute features | Acute bronchitis | NA (binary) | ICD-10: J20.*, J40 and ICD-9: 466.0, 490 |
|  | Anorexia | NA (binary) | ICD-10: R63.0, R63.8 and ICD-9: 783.0, 783.9 |
|  | Body temperature | °C | LOINC: 8310-5 |
|  | Confusion | NA (binary) | ICD-10: R41.0, R41.82 and ICD-9: 780.97 |
|  | Cough | NA (binary) | ICD-10: R05 and ICD-9: 786.2 |
|  | Diarrhea | NA (binary) | ICD-10: R19.7 and ICD-9: 787.91 |
|  | Fatigue | NA (binary) | ICD-10: R53.1, R53.81, R53.83 and ICD-9: 780.79 |
|  | Fever | NA (binary) | ICD-10: R50.9 and ICD-9: 780.60 |
|  | Headache | NA (binary) | ICD-10: R51 and ICD-9: 784.0 |
|  | Hemoptysis | NA (binary) | ICD-10: R04.2 and ICD-9: 786.30 |
|  | Hospitalization (inpatient) | NA (binary) | Considered if reported admission–discharge period overlapping with extraction time window |
|  | Myalgia | NA (binary) | ICD-10: M79.1, M79.10, M79.11, M79.12, M79.18 and ICD-9: 729.1 |
|  | Pneumonia | NA (binary) | ICD-10: J12.*, J13, J14, J15.*, J16.*, J17, J18.* and ICD-9: 480.*, 481, 482.*, 483.*, 484.*, 485, 486, 487.0, 488.01, 488.11, 488.81 |
|  | Rhinorrhea | NA (binary) | ICD-10: J34.89 and ICD-9: 478.19 |
|  | Shortness of breath | NA (binary) | ICD-10: R06.02 and ICD-9: 786.05 |
|  | Sore throat | NA (binary) | ICD-10: J02.9 and ICD-9: 462 |
|  | Sputum | NA (binary) | ICD-10: R09.3 and ICD-9: 786.4 |
|  | Vomiting | NA (binary) | ICD-10: R11.10 and ICD-9: 536.2, 787.03 |
| Chronic features | Active smoking | NA (binary) | Based on reported habit |
|  | Asthma | NA (binary) | ICD-10: J45.* and ICD-9: 493.* |
|  | BMI | kg/m<sup>2</sup> | LOINC: 39156-5, or computed from weight (29463-7) and height (8302-2) |
|  | Cardiovascular disease | NA (binary) | ICD-10: I20.*, I21.*, I25.*, I48.*, I50.*, I63.*, I65.*, I67.*, I73.* and ICD-9: 410.*, 412.*, 413.*, 414.*, 427.*, 428.*, 429.*, 433.*, 434.*, 437.*, 443.* |
|  | Chronic kidney disease | NA (binary) | ICD-10: E10.21, E10.22, E10.29, E11.21, E11.22, E11.29, I12.0, I12.9, I13.0, I13.10, I13.11, I13.2, N04.*, N05.*, N08, N18.*, N19, N25.9 and ICD-9: 250.40, 250.41, 250.42, 250.43, 403.*, 404.*, 581.81, 581.9, 583.89, 585.*, 588.9 |
|  | Chronic obstructive pulmonary disease | NA (binary) | ICD-10: J44.* and ICD-9: 491.*, 493.2* |
|  | Diabetes | NA (binary) | ICD-10: E10.*, E11.*, E13.* and ICD-9: 250.* |
|  | Hypertension | NA (binary) | ICD-10: I10, I15.* and ICD-9: 401.*, 405.* |
|  | Immunodeficiency | NA (binary) | ICD-10: B20, D80.*, D81.*, D82.*, D83.*, D84.*, D86.*, D89.* and ICD-9: 042, 279.* |
|  | Nicotine dependence | NA (binary) | ICD-10: F17.* and ICD-9: 305.1 |
|  | Obesity | NA (binary) | ICD-10: E66.0*, E66.1, E66.2, E66.8, E66.9 and ICD-9: 278.00, 278.01, 278.03 |
|  | Paralytic syndromes | NA (binary) | ICD-10: G80.*, G81.*, G82.*, G83.* and ICD-9: 342.*, 343.*, 344.* |

\* symbolizes a wildcard for ICD subcategory codes.

## 3 Discussion

In this work, a prognostic model was created based on real-world data from 12652 patients to predict at COVID-19 diagnosis, whether patients will enter a critical state within the next 28 days or not. In addition to demographic, and clinical data, hospitalization and insurance types were used as predictors. Our results based on new 3164 patients unseen during training showed high predictive performance (sensitivity of 0.841 and specificity of 0.762) and well-calibrated output probabilities with a minor tendency to over-forecast probabilities above 0.5. Furthermore, the interpretability analysis identified older age, pneumonia, higher BMI, diabetes, male gender, shortness of breath, and cardiovascular disease as most important predictive factors for critical state.

### 3.1 Validity of the COVID-19 dataset

Around 16 000 US patients diagnosed with COVID-19 met the inclusion criteria. To the best of our knowledge, it is one of the largest cohorts used for COVID-19 progression modeling to date based on EHR data.

The definitions used for severe state or critical state vary across different sources (e.g., intubation prior to ICU admission, discharge to hospice, or death ^[17]^, moderate to severe respiratory failure ^[11]^, oxygen requirement greater than 10L/min or death ^[13]^), or are not described in detail. Based on the definition by the World Health Organization ^[24]^ including sepsis, septic shock, and respiratory failure (e.g., acute respiratory distress syndrome (ARDS)), the proportion of patients entering critical state (13.0%) in our study is within the range of prevalence (12.6% to 23.5%) reported in a review covering 21 studies ^[25]^.

Similarly, case fatality rates vary across US states and countries, as they directly depend on factors such as the number of tested people, demographics, socioeconomics, or healthcare system capacities. The death rate for the entire US is estimated to be 3% ^[2]^ (status August 26, 2020). In the present work, the reported proportion of people assumed to be deceased because of COVID-19 is 3.4%. These differences may be justified in part by the fact that in these sources the outcome (i.e., potential death) of recent cases is yet unknown when computing the case fatality rate, hence leading to underestimation. As our analysis enforces at least 7 weeks of data after diagnosis date increasing changes of knowing the patients’ outcomes, this underestimation can be reduced. Nevertheless, death is not reliably reported in EHRs and records were de-identified making linking to public death records not feasible.

Regarding demographics of our cohort, there are only minor dissimilarities to numbers reported by the Centers for Disease Control and Prevention (CDC) or US states. The interquartile range of the age distribution of our cohort (32–63 years) matches with the 33-63 years for COVID-19 cases across the entire US ^[26]^. The racial breakdown varies strongly across different US states. Given that Explorys clients are mostly in metropolitan areas, there is a higher proportion of African Americans in the present EHR dataset compared to US average ^[27]^. As Caucasian and African American together represent 92.7% of the dataset, there is a strong negative correlation between the two features, for which reason the majority group (race (Caucasian)) was considered as baseline and removed from the feature set. The proportion of female cases (56.8%) is more pronounced compared to the US-wide incidences of 406 (female) and 401 (male) cases per 100000 persons also showing a marginally higher rate for females than males, respectively ^[26]^. Since the medical system captured by Explorys is separate to the billing system, it can be expected that information on insurance types is not widely available. As a matter of fact, less than 10% of patients have a reported insurance type.

The most common underlying comorbidities identified through ICD codes in our cohort are hypertension, obesity, cardiovascular disease, diabetes, and chronic lung disease (includes asthma and chronic obstructive pulmonary disease). As this is in line with statistics from the CDC ^[26,28]^ as well as other studies conducted in China (e.g., ^[29]^) and the prevalence of such features is not affected by any time window restrictions (i.e., the entire patient history was considered), it substantiates the validity of the Explorys data.

Since the aim of the present work is to develop a model for predictions at the time point of COVID-19 diagnosis, symptoms identified through ICD codes (e.g., fever or cough) are only extracted from the 14 days previous to the COVID-19 diagnosis. As the COVID-19 diagnosis may be early or late in the disease progression, there is the possibility to capture either early or late symptoms depending on each case. However, due to the time window restriction, the prevalence of reported symptoms tends to be lower compared to statistics including reported symptoms during the entire course of the disease ^[26]^. Moreover, outpatient symptoms based on ICD codes may be under-documented, as hospitals may not get paid for their diagnosis. Despite these lower numbers, the most common symptoms in our cohort, namely cough, fever, and shortness of breath, are confirmed by other reports and studies ^[26,30,31]^.

Overall, the size and quality of the EHR dataset based on the Explorys database demonstrates high value with regards to demographics, chronic features, and acute symptoms, despite its sparsity.

### 3.2 Performance

Although our dataset is based on sparse real-world data, our prognostic model shows an excellent model performance in terms of ROC AUC (0.872 [0.868, 0.877]) ^[32]^ and a substantial improvement of the PR AUC (0.500 [0.488, 0.509]) compared to chance level (0.130). Optimizing the decision threshold by maximizing the Youden’s J statistic lead to a sensitivity of 0.841 [0.819, 0.856] and a specificity of 0.762 [0.740, 0.781]. Depending on the medical requirements for the prognostic model in terms of sensitivity and specificity, the threshold could easily be adjusted for a real application. As different types of datasets, inclusion/exclusion criteria, features, and prediction target definitions were used in other papers presenting the development of models predicting COVID-19 critical state, (e.g., ^[13,17]^, or review ^[8]^), it renders it difficult to do a direct performance comparison (reported metrics were in the following ranges: ROC AUC 0.81–0.99, PR AUC 0.56–0.71, sensitivity 0.70–0.94, specificity 0.75–0.85). Furthermore, some publications do not mention metrics (e.g., PR AUC, or sensitivity and specificity) required to properly evaluate performance on an imbalanced dataset, which is the case for this type of COVID-19 prognosis. Unlike other papers ^[11,13,17]^ usually performing a cross-validation or using a limited number of independent sets for the testing, the present approach used random, stratified train-test splits repeated 100 times to obtain a distribution of performance. Such an approach has the advantage of providing a better understanding of the generalizability of the model and the robustness of the performance estimate, as it is likely that a single test set might underestimate or overestimate the real performance for small testing sets. Even though our model was trained on data coming from many hospitals compared to other work being only based on a single or limited number of contributors, an external validation should be performed to better assess its generalizability.

Most publications on prognosis prediction models do not report model calibration ^[8]^, with the exception of a few ^[13,18]^. The present model based on the Explorys dataset is well-calibrated, showing only minor tendency to over-forecast probabilities above 0.5. We hypothesize that this over-forecast comes from the fact that features encoding care or treatments (e.g., drugs) were not included in the model. Assuming that treatments reduce the probability of entering critical state, taking a treatment will lead to an overestimated probability by the model, as this information is not available to the model. In any case, over-forecast accentuating cases with relatively high probability is preferable to under-forecast, where patients with high probability of critical case may not be identified.

Overall, our prognostic model shows excellent performance and has the advantage to provide a calibrated risk score instead of a binary classification. This could potentially help healthcare professionals to create a more fine-graded risk stratification of patients.

### 3.3 Model interpretability

Pneumonia appeared among the top features, as pneumonia is a diagnosis defining moderate and severe cases ^[24]^, which are precursor stages for critical state due to COVID-19 disease. The results from a study with 1099 patients showed that patients with severe disease had a higher incidence of physician-diagnosed pneumonia than those with non-severe disease ^[33]^.

As identified by the interpretability analysis, older age is an important risk factor. This has been confirmed by many studies showing its relevance in progressing to grade IV and V on the pneumonia severity index and mortality of COVID-19 patients ^[34–36]^. The developed model was also able to endorse existing results showing that men are, despite similar prevalence to women, more at risk for worse disease severity, independent of age ^[37]^. Similarly, obesity has been reported as a factor increasing probability of higher disease severity and lethality ^[33,38,39]^. While according to our interpretability analysis the feature obesity shows marginal importance in the output of the model, the feature BMI is among the top features leading to high risk (in case of high BMI). It can be assumed that the feature obesity with a prevalence of 25.1% in our dataset compared to age-adjusted prevalence of obesity in the US is around 35% ^[40]^ is under-reported in the EHR data of our cohort. The median BMI in our dataset is very close to the threshold from overweight to obesity (BMI of > 30 kg/m^2^). Hence it can be concluded that approximately 50% of our patients are obese. In addition, the BMI feature is a continuous variable with only 16.7% missing entries, having thus more information content and, as a result, shows higher predictive importance than obesity.

In line with the literature, the following comorbidities were also shown to drive high probabilities for critical state: diabetes ^[41–43]^, chronic kidney disease ^[44–46]^, and cardiovascular diseases ^[36,47,48]^. As a matter of fact, many elderly patients with these comorbidities use Angiotensin-converting enzyme (ACE) inhibitors and Angiotensin-receptor blockers (ARBs) which up-regulate the ACE-2 receptor ^[49]^. Given that ACE-2 receptor has been proposed as a functional receptor for the cell entry mechanism of coronaviruses, it has been hypothesized that as a consequence this may lead to a higher prevalence and elevated risk for a severe disease progression after SARS-CoV-2 infection ^[50]^.

Our model also revealed disparities in terms of probability for critical state between races: African American seem to have a higher risk of entering critical state compared to Caucasian. This fact has been verified in several states, among others Louisiana where around 70% of deaths have occurred among African Americans, although they represent only one third of the state’s population ^[51]^. While a higher prevalence of comorbidities such as hypertension, diabetes, obesity, and cardiovascular disease among African Americans may be one reason for these disproportion, also late lockdowns in southern states or social determinants (e.g., living in poor areas with high housing density, high crime rates, poor access to healthy foods) may be strong contributors ^[51,52]^. For ethnicity, on the other hand, consistent effect can be concluded based on SHAP values for Hispanics versus non-Hispanics. While socioeconomic factors and insurance types may also have an influence on the probability of severe disease progression, the SHAP analysis did not reveal any major trends. However, **Figure 5** shows that privately insured patients may have a marginally lower risk for critical state, as they may be able to seek earlier and better care. As there there are no “selfpay” patients, who may be more reluctant to seek early medical care due to costs, in our cohort, this hypothesis can unfortunately not be supplemented with additional evidence.

The two primary symptoms influencing the progression of the disease based on the present analysis are shortness of breath (dyspnea) and cough, both prevalent symptoms for COVID-19 ^[31]^. Interestingly, they have opposite effects on the prediction probability of the model, with shortness of breath increasing and cough decreasing the probability for critical state. This can be explained by the fact that cough is an early symptom during mild or moderate disease, and shortness of breath develops in the late course of illness. This concurs with statistical reports from China showing higher prevalence of shortness of breath in severe cases and a higher prevalence of cough in non-severe cases and survivors ^[29,53,54]^. Hence, if cough is reported, this may indicate that the disease is still in early stage and there is the chance that it may not lead to a critical state, whereas if shortness of breath is reported, chances for further disease progression may be much higher. Furthermore, hospitals may not report outpatient symptoms such as cough, whereas they may report more critical symptoms such as shortness of breath more reliably. This means that it is highly likely that many of the patients in our cohort without an ICD code entry for cough actually may have had cough, in particular given that it is a highly prevalent symptom. This may considerably contribute to this rather surprising result. Fever may be at the same time an early appearing symptom but has also been shown to be developed later during hospitalization ^[31,33]^. In addition, the same prevalence of fever has been reported in survivors and non-survivors ^[29]^. This may also explain why it is more difficult to use it as a predictive feature, unlike for example cough, despite being also among the most prevalent symptoms ^[30]^. Nonspecific neurological symptoms like headache and confusion are less commonly reported ^[30]^. Nevertheless, the right plot in **Figure 5** reveals that the presence of confusion significantly contributes to an increase in the model’s output probability, despite having low overall importance (which in turn is also driven by the low prevalence within our dataset). While headaches may have many potential origins not necessarily related to COVID-19, confusion may be a clearer precursor of neuroinvasion of SARS-CoV-2, which has been suggested to potentially lead to respiratory failure ^[55]^.

Overall, the findings of this work are in line with results from the vast number of studies reported in the literature and the interpretability analysis provides evidence for the validity of the prognostic prediction modeling.

### 3.4 Limitations

EHRs can be a powerful data source to create evidence based on real-world data, especially when combined with a platform facilitating the structured extraction of data. However, there are trade-offs to be made when doing analyses on EHR data in contrast to the analysis of clinical study data ^[56]^. One major limitation is that patients may get diagnoses, treatments, or observations outside of the hospital network covered by Explorys, resulting in sparse patient histories. Other challenges are potential over-and under-reporting of diagnoses, observations, or procedures. For example, clinicians may enter an ICD-10 code for COVID-19 when ordering a SARS-CoV-2 test leading to over-documentation and “false positive” entries. On the other hand, replying only on test results may increase the risk that tested patients only performed the test at a hospital with the Explorys network, but did not get diagnosed and treated within the same hospital, which would lead to potentially “false negatives” in terms of target labeling. For this reason the inclusion criteria for our cohort was based on the combination of an ICD code entry for COVID-19 with a positive SARS-CoV-2 test result, to increase the probability of only including patients with actual COVID-19. This highly sparse data may also require imputation, as there is rarely a patient with a complete data record, especially when the set of features is large. The method of imputation may also introduce additional biases which are difficult to control. As particularly important in predictive modeling, it was ensured that the imputation was based purely on the train set to avoid additional information leakage. Furthermore, to ensure data privacy and prevent re-identification, patients’ age is truncated, and death dates and related diagnoses and procedures are not available in Explorys data. As the latter is highly relevant for the present modeling, several assumptions had to be taken. Nevertheless, resulting death rates correspond well to official COVID-19-related death rates in the US or relevant states.

An additional limitation and potential bias is linked to the data extraction using time windows. Even though the window lengths were motivated by medical reasoning, they are subject to trade-offs which is not the case for clinical studies due to precise protocols: extending the windows to capture enough information spread over multiple visits and account for delays in EHR entries, versus remaining recent enough and related to COVID-19. Furthermore, the features used in this model do not capture the time information for the individual samples (e.g., how many days before COVID-19 diagnosis the ICD code for fever was entered into the system). In addition, it could be that the reference for the time windows is not accurate, as the ICD code or LOINC entry used as COVID-19 diagnosis proxy may not have been the actual first diagnosis of the patient.

The model was based on US data from hospitals of the Explorys network, sampling mostly metropolitan areas. This resulted for example in a higher ratio of African Americans compared to the US average, it is highly likely that there are demographic and socioeconomic biases, in addition to the fact that economically disadvantaged patients may seek medical help too late. Moreover, the data reflects the American healthcare system in terms or testing, diagnosing, and treating procedures as well as reporting.

Despite these limitations, RWE can retrospectively generate insights on a scale which would not be feasible with an observational clinical study. Thus, it may be a starting point for subsequent, more focused clinical studies. Furthermore, approaches based on RWE might even have higher clinical applicability due to their incorporation of statistical noise while model training ^[57]^.

### 3.5 Conclusions

The results of this work demonstrate that it is possible to develop an explainable machine learning model based on patient-level EHR data to predict at the time point of COVID-19 diagnosis whether individual patients will progress into critical state in the following four weeks. Without the necessity of relying on multiple laboratory test results or imaging such as computer tomography (CT), this model holds promise of clinical utility due to the simplicity of the relevant features and its adequate sensitivity and specificity. Even though this prognostic model for critical state has been trained and evaluated on one of the largest COVID-19 cohorts to date with EHR data from around 16 000 patients, it includes predominantly cases from metropolitan areas within the US and may therefore be biased towards sub-populations of the US and the American healthcare system. To prove its generalizability before being considered for clinical implementation, it should be validated with other datasets. Such RWE models have the potential to identify new risk factors by mining EHRs. This model could also be augmented with treatment features (e.g., drugs or other interventions) after diagnosis in order to predict whether the respective treatments would lead to an improvement (i.e., reduction of the probability of entering critical state). RWE approaches will never replace clinical studies to validate risk factors or evaluate treatment effectiveness. Nevertheless, these types of retrospective real-world data analyses can support other research generally requiring much higher efforts and costs: They could help identifying responder groups or informing the design of clinical trials, with the aim of making research more efficient and accelerating the avenue to personalized treatment and eventually reduced burden on the healthcare system.

## 4 Methods

### 4.1 RWE Insights Platform

This work was achieved by using the *RWE Insights Platform*, a data science platform for analyses of medical real-world data to generate RWE recently developed by IBM. The *RWE Insights Platform* is a data science pipeline facilitating the setup, execution, and reporting of analyses of medical real-world data to discover RWE insights in an accelerated way. The platform architecture is built in a fully modular way to be scalable to include different types of analyses (e.g., treatment pathway analysis, treatment response predictor analysis, comorbidity development analysis) and interface with different data sources (e.g., the Explorys database).

For the present use case of COVID-19 prognosis prediction, we used the comorbidity development analysis which allows defining a cohort, an outcome to be predicted, a set of predictors, and relative time windows for the extraction of the samples from the data source. New data-extraction modules for specific disease, outcome, treatments, and variables for the current use case were developed.

The *RWE Insights Platform* has been developed using open-source tools and includes a front end based on HTML and CSS interfacing via a Flask RESTful API to a Python back end (python 3.6.7) using the following main libraries: imbalanced-learn 0.6.2, numpy 1.15.4, pandas 0.23.4, scikit-learn 0.20.1, scipy 1.1.0, shap 0.35.0, statsmodel 0.90.0, and xgboost 0.90. The platform is a proprietary software owned by IBM. The detailed description of the *RWE Insights Platform* is beyond the scope of this publication.

### 4.2 Real-world data source

Our work was based on de-identified data from the Explorys database. The Explorys database is one of the largest clinical datasets in the world containing EHRs of around 64 million patients across more than 360 hospitals in the US ^[58]^. Data were standardised and normalised using common ontologies, searchable through a Health Insurance Portability and Accountability Act (HIPAA)-enabled, de-identified dataset from IBM Explorys. Individuals were seen in multiple primary and secondary healthcare systems from 1999 to 2020 with a combination of data from clinical electronic medical records, health-care system outgoing bills, and adjudicated payer claims. The de-identified EHR data include patient demographics, diagnoses, procedures, prescribed drugs, vitals, and laboratory test results. Hundreds of billions of clinical, operational, and financial data elements are processed, mapped, and classified into common standards (e.g., ICD, SNOMED, LOINC, and RxNorm) within the data lake. The Explorys database has been proven to be useful in many retrospective data analyses for different applications (e.g., refs. ^[59–63]^). As data in Explorys is updated continuously, a view of the database was created and frozen on August 26, 2020 for reproducibility of this work.

### 4.3 Cohort

The cohort included all patients in the Explorys database having a documented diagnosis of COVID-19 and a reported positive entry for a SARS-CoV-2 test, both since December 1, 2019. As the new ICD-10 code U07.1 for COVID-19 cases confirmed by laboratory testing has been created and pre-released a couple of months after pandemic onset, already existing ICD codes related to coronavirus (B34.2 Coronavirus infection, unspecified and B97.29 Other coronavirus as the cause of diseases classified elsewhere) were also included, as hospitals may have used them for early cases. Based on their appearance in Explorys, the following LOINC codes for the SARS-CoV-2 tests were included: 94309-2, 94500-6, 94502-2, 94505-5, 94507-1, and 94547-7 ^[64]^. The December 2019 cutoff was instituted to be consistent with the spread of COVID-19 in the US and to limit inclusion of patients who may have been diagnosed with other forms of coronavirus besides SARS-CoV-2. In case of multiple entries per patient after December 1, 2019, the first ICD code or LOINC entry date was used as COVID-19 diagnosis date. In order to have enough data to extract the patient’s outcome, the diagnosis date had to be at least 7 weeks before the freeze date of the database (August 26, 2020), as it may take up to 7 weeks from symptom onset to death ^[65]^.

### 4.4 Prediction target

Critical state was used as a binary prediction target and included sepsis, septic shock, and respiratory failure (e.g., ARDS) ^[24]^. Severe sepsis is associated with multiple organ dysfunction syndrome. The precise definition based on ICD codes used for critical state is listed in **Table 2**. In case of multiple entries for a patient, the first entry was retained. In addition, the date of the entry for critical state had to be in a window of [0, +28] days (boundaries included) after the diagnosis date to be eligible, as illustrated in **Figure 7**). Four weeks were chosen to ensure coverage of the majority of critical outcomes, as the interquartile range of time from illness onset to sepsis and ARDS were reported to be [7, 13] and [8, 15] days, respectively ^[29]^. Patients with an eligible entry for critical state were labeled as entering critical state, whereas patients eligible based on cohort definitions without any entry for critical state were labeled as not entering critical state. One exception to these rules were patients who are flagged as deceased in the Explorys database. In order to include death cases potentially related to COVID-19 in the critical state group, and as death dates and records with diagnoses and procedures relating to the patient’s death are not available in the Explorys data to avoid re-identification of patients and ensure data privacy, patients with one of the following conditions were also labeled as entering critical state: deceased with an entry for critical state within the window, deceased with an entry for critical state within and after the window, or deceased without any entry for critical state (and thus excluding deceased patients with an entry for critical state before the window). In the latter case, the date was set to the end of the window for critical state entries. To validate these assumptions, the proportion of patients assumed to be deceased due to COVID-19 in our cohort was compared to epidemiological numbers.

**Figure 7.**
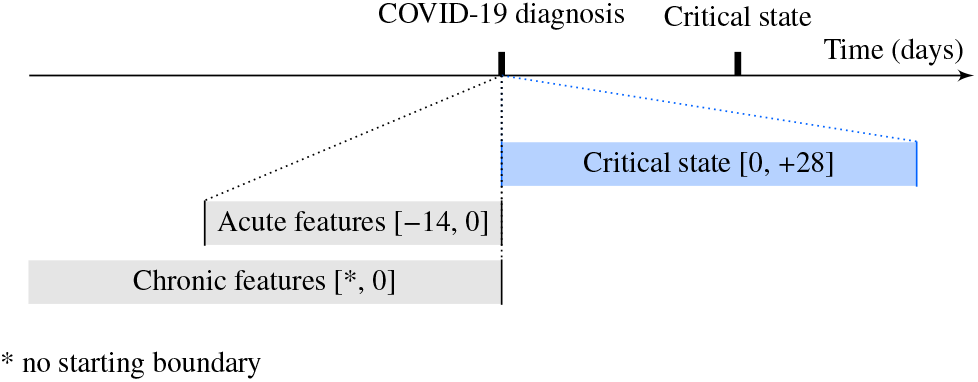
Time windows for prediction target and feature extraction. Schematic illustration of time window definitions relative to the COVID-19 diagnosis or to the critical state (time not to scale). The brackets define the boundaries (included) in days.

### 4.5 Features

Features were mainly grouped into “acute” features and “chronic” features. Acute features are a set of features which should be temporally close to the COVID-19 diagnosis (e.g., body temperature, symptoms potentially related to COVID-19, or hospitalization prior to the diagnosis), whereas chronic features are a set of features which have no direct temporal relation to the COVID-19 diagnosis (e.g., chronic comorbidities, measurable demographics, or long-term habits). Features were selected based on potential risk factors and predictors related to COVID-19 reported in the literature. **Figure 7** illustrates their difference in terms of time windows for extraction. A negative value for boundaries of time window definitions stand for dates prior to the reference date (e.g., prior to the diagnosis date). Ideally, acute features should have been recorded for higher consistency at diagnosis date. However, this may not be always the case in the EHR compared to data from clinical studies. To account for recorded symptoms previous to the diagnosis (e.g., through telemedicine before performing a SARS-CoV-2 test or due to potentially required multiple testing because of false negatives delaying diagnosis), a time window of [–14, 0] days before the diagnosis was used to extract acute features. Patients were considered hospitalized (inpatient) if the reported admission-discharge period of the hospitalization overlapped with the acute feature extraction time window. Entries for chronic features were considered if prior to the diagnosis date, without additional restriction. Demographic features which were not restricted to any time window (e.g., gender or race) or required a special way of extraction/computation (e.g., age) are grouped as “special” features and are not represented in **Figure 7**. As part of the de-identification process, for patients over 90 years of age, the age is truncated to 90 years. Similarly, the age of all patients born within the last 356 days is be set to 0 years. The full list of features including their definitions (e.g., based on ICD or LOINC codes) is provided in **Table 3**, grouped by extraction time window type. As features entries (especially relevant for chronic features) may have been entered several years ago, ICD-9 codes were used as well for the extraction. In general, the last entry within the specific extraction time window was used to construct the feature, except if described otherwise in **Table 3**.

### 4.6 Dataset preparation and modeling approach

The full dataset was constructed based on COVID-19 diagnosis including binary prediction target labels for critical state and enriched by the various features. Patients with missing age or gender information were removed from the dataset, and all missing binary features (i.e., obtained from ICD code entries) of **Table 3** were imputed with zero. Descriptive distribution statistics were created for all features, and features with more than 90% missing values were removed from the feature set. For the remaining feature set, the concurvity (non-linear collinearity) among features was assessed using Kendall’s *τ*, a non-parametric measure of correlation. In case of |*τ*| > 0.7 ^[66]^, the feature with more missing values was removed from the feature set. In case of equal number of missing values, the feature with the higher mean was removed in order to keep the minorities and make the larger group part of the predicted probability baseline. To train and evaluate the model, the dataset was split into a train set (80%) and test set (20%) using stratification of the prediction target. This procedure was repeated 100 times based on different random seeds to get a distribution and confidence intervals of the model performance and feature importance, as performance may change depending on the choice of splits.

For each random split the following steps were executed: The non-binary features of the train set and the test set were imputed based on the feature medians of the train set to avoid data leaking. An XGBoost model was trained on the train set using default parameters of the XGBoost Python package without additional hyperparameter tuning. XGBoost is a decision-tree-based ensemble machine learning algorithm using a gradient boosting framework. Gradient tree boosting models have shown to outperform other types of models on a large set of benchmarking datasets ^[67]^. The trained XGBoost model was subsequently used to create predictions for the test set.

### 4.7 Performance analysis and model interpretability

The performance of the model was evaluated on the test set for each random train-test split seed and reported with median and interquartile range across seeds. This provides a distribution of expected performance, if a new model would be trained on similar data. Following metrics were computed: receiver operating characteristic (ROC) curve and precision recall (PR) curve as well as their respective areas under the curve (ROC AUC and PR AUC). The confusion matrix, sensitivity, and specificity were reported for the optimal probability classification threshold. This threshold was obtained based on maximizing the largest Youden’s J statistic (corresponding to the largest geometric mean as a metric for imbalanced classification seeking for a balance between sensitivity and specificity). Furthermore, the calibration of the model was reported, comparing binned mean predicted values (i.e., probabilities) to the actual fraction of positives (labeled as critical state) ^[68]^, in order to evaluate whether the predicted probability is realistic and can provide some confidence on the prediction.

Interpretability of the model was generated using Tree SHAP ^[23]^, a version of SHAP (SHapley Additive exPlanations) optimized for tree-based models. SHAP is a framework to explain the contribution of feature values to the output of individual predictions by any type of model and to compute the global importance of features. This individual contribution is expressed as SHAP value, corresponding to log-odds (output of the trees in XGBoost), before they are converted into probabilities with a logistic function. The global feature importance as well as a summary plot of individual contributions including feature values were created. In our case, a positive SHAP value indicates a contribution towards increased probability for critical state, whereas a negative SHAP value indicates a reduction of probability for critical state.

## Data Availability

The patient data that support the findings of this study are available from IBM Explorys but restrictions apply to the availability of these data, which were used under license for the current study, and so are not publicly available.

## Acknowledgments

The authors would like to thank Tobias Egli, Oliver Müller, Ajandek Peak, and Sylvia Schumacher for contributing to the development of the *RWE Insights Platform*, and in particular Tobias Egli and Oliver Müller for their feedback on the manuscript. Further thanks go to the IBM Watson Health® team for enabling this project by providing access to the Explorys dataset, Ben Kolt for support and advice related to EHR data as well as Brenna Brady and Anil Jain, MD, VP and Chief Health Information Officer, IBM Watson Health, for critical review of the manuscript. The RWE *Insights Platform* project is supported and sponsored by Paolo Bassignana and Lars Böhm (IBM Switzerland Ltd).

## Author contributions

M.R. and Y.K. lead the development of the *RWE Insights Platform*, contributed to the conception of this work, developed the methodology, implemented the use case and the modeling approach, performed the analysis, interpreted the results, and drafted the manuscript. Both authors revised the manuscript and approved the final version.

## Competing interests

M.R. and Y.K. are employees of IBM Switzerland Ltd.

## Code availability

Custom codes were made for the analysis using open source libraries (python 3.6.7, numpy 1.15.4, pandas 0.23.4, scikit-learn 0.20.1, scipy 1.1.0, shap 0.35.0, and xgboost 0.90). The custom codes are owned by IBM and cannot be shared for proprietary reasons.

## Notes

### Funding Statement

No third party funding was received for this work.

### Author Declarations

The project was approved by the Data Access and Control Board (IBM Watson Health) to further research into COVID-19 for the greater good. Patients in the US opt in by default and need to actively opt out. If the opt out, their data will not be part of the Explorys database used in this work.

## References

[1] Gorbalenya, A. E. et al. The species severe acute respiratory syndrome-related coronavirus: classifying 2019-nCoV and naming it SARS-CoV-2. Nature Microbiology 5, 536–544 (2020).

[2] Johns Hopkins University (JHU). COVID-19 dashboard by the Center for Systems Science and Engineering (CSSE) at Johns Hopkins University (JHU) (2020). Accessed on 2020-08-26 https://coronavirus.jhu.edu/map.html.

[3] Peeri, N. C. et al. The SARS, MERS and novel coronavirus (COVID-19) epidemics, the newest and biggest global health threats: what lessons have we learned? International Journal of Epidemiology (2020).

[4] Anderson, R. M., Heesterbeek, H., Klinkenberg, D. & Hollingsworth, T. D. How will country-based mitigation measures influence the course of the COVID-19 epidemic? The Lancet 395, 931–934 (2020).

[5] Armocida, B., Formenti, B., Ussai, S., Palestra, F. & Missoni, E. The italian health system and the COVID-19 challenge. The Lancet Public Health 5, e253 (2020).

[6] Ranney, M. L., Griffeth, V. & Jha, A. K. Critical supply shortages — the need for ventilators and personal protective equipment during the Covid-19 pandemic. New England Journal of Medicine 382, e41 (2020).

[7] Bullock, J. et al. Mapping the landscape of artificial intelligence applications against COVID-19. *arXiv* (2020). Preprint at https://arxiv.org/abs/2003.11336, 2003.11336.

[8] Wynants, L. et al. Prediction models for diagnosis and prognosis of COVID-19 infection: systematic review and critical appraisal. BMJ 369 (2020).

[9] Bai, X. et al. Predicting COVID-19 malignant progression with AI techniques. *medRxiv* (2020). Preprint at https://www.medrxiv.org/content/10.1101/2020.03.20.20037325v2.

[10] Feng, Z. et al. Early prediction of disease progression in 2019 novel coronavirus pneumonia patients outside wuhan with CT and clinical characteristics. medRxiv (2020). Preprint at https://www.medrxiv.org/content/10.1101/2020.02.19.20025296v1.

[11] Ferrari, D. et al. Machine learning in predicting respiratory failure in patients with COVID-19 pneumonia - challenges, strengths, and opportunities in a global health emergency. medRxiv (2020). Preprint at https://www.medrxiv.org/content/10.1101/2020.05.30.20107888v2.

[12] Gong, J. et al. A tool to early predict severe 2019-novel coronavirus pneumonia (COVID-19): A multicenter study using the risk nomogram in Wuhan and Guangdong, China. medRxiv (2020). Preprint at https://www.medrxiv.org/content/10.1101/2020.03.17.20037515v2.

[13] Haimovich, A. et al. Development and validation of the COVID-19 severity index (CSI): a prognostic tool for early respiratory decompensation. *medRxiv* (2020). Preprint at https://www.medrxiv.org/content/10.1101/2020.05.07.20094573v2.

[14] Jiang, X. et al. Towards an artificial intelligence framework for data-driven prediction of coronavirus clinical severity. Computers, Materials & Continua 63, 537–551 (2020).

[15] Liu, J. et al. Neutrophil-to-lymphocyte ratio predicts severe illness patients with 2019 novel coronavirus in the early stage. *medRxiv* (2020). Preprint at https://www.medrxiv.org/content/10.1101/2020.02.10.20021584v1.

[16] Petrilli, C. M. et al. Factors associated with hospitalization and critical illness among 4,103 patients with COVID-19 disease in New York City. *medRxiv* (2020). Preprint at https://www.medrxiv.org/content/10.1101/2020.04.08.20057794v1.

[17] Vaid, A. et al. Machine learning to predict mortality and critical events in COVID-19 positive New York City patients. *medRxiv* (2020). Preprint at https://www.medrxiv.org/content/10.1101/2020.04.26.20073411v1.

[18] Xie, J. et al. Development and external validation of a prognostic multivariable model on admission for hospitalized patients with COVID-19. medRxiv (2020). Preprint at https://www.medrxiv.org/content/10.1101/2020.03.28.20045997v2.

[19] Yan, L. et al. A machine learning-based model for survival prediction in patients with severe COVID-19 infection. *medRxiv* (2020). Preprint at https://www.medrxiv.org/content/10.1101/2020.02.27.20028027v3.

[20] DeCaprio, D. et al. Building a COVID-19 vulnerability index. *arXiv* (2020). Preprint at https://arxiv.org/abs/2003.07347, 2003.07347.

[21] Benchimol, E. I. et al. The reporting of studies conducted using observational routinely-collected health data (RECORD) statement. PLOS Medicine 12, e1001885 (2015).

[22] Collins, G. S., Reitsma, J. B., Altman, D. G. & Moons, K. G. Transparent reporting of a multivariable prediction model for individual prognosis or diagnosis (TRIPOD): The TRIPOD statement. Annals of Internal Medicine 162, 55–63 (2015).

[23] Lundberg, S. M. et al. From local explanations to global understanding with explainable ai for trees. Nature Machine Intelligence 2, 56–67 (2020).

[24] WHO. Severe acute respiratory infections treatment centre. Tech. Rep., Avenue Appia 20, 1202 Geneva, Switzerland (2020).

[25] Hu, Y. et al. Prevalence and severity of corona virus disease 2019 (COVID-19): A systematic review and meta-analysis. Journal of Clinical Virology 127, 104371 (2020).

[26] Stokes, E. K. et al. Coronavirus disease 2019 case surveillance—United States, January 22–May 30, 2020. MMWR Morb Mortal Wkly Rep 69, 759–765 (2020).

[27] Census Bureau, U. U.S. Census Bureau QuickFacts: United States (2020).

[28] Garg, S. et al. Hospitalization rates and characteristics of patients hospitalized with laboratory-confirmed coronavirus disease 2019 — COVID-NET, 14 states, march 1–30, 2020. MMWR Morb Mortal Wkly Rep 69, 458–464 (2020).

[29] Zhou, F. et al. Clinical course and risk factors for mortality of adult inpatients with COVID-19 in Wuhan, China: a retrospective cohort study. The Lancet 395, 1054 – 1062 (2020).

[30] Chen, N. et al. Epidemiological and clinical characteristics of 99 cases of 2019 novel coronavirus pneumonia in wuhan, china: a descriptive study. The Lancet 395, 507–513 (2020).

[31] Yang, X. et al. Clinical course and outcomes of critically ill patients with SARS-CoV-2 pneumonia in Wuhan, China: a single-centered, retrospective, observational study. The Lancet Respiratory Medicine 8, 475–481 (2020).

[32] Mandrekar, J. N. Receiver operating characteristic curve in diagnostic test assessment. Journal of Thoracic Oncology 5, 1315–1316(2010).

[33] Guan, W.-j. et al. Clinical characteristics of coronavirus disease 2019 in China. New England Journal of Medicine 382, 1708–1720 (2020).

[34] Du, R.-H. et al. Predictors of mortality for patients with COVID-19 pneumonia caused by SARS-CoV-2: a prospective cohort study. European Respiratory Journal 55 (2020).

[35] Liu, K., Chen, Y., Lin, R. & Han, K. Clinical features of COVID-19 in elderly patients: A comparison with young and middle-aged patients. Journal of Infection 80, e14–e18 (2020).

[36] Mehra, M. R., Desai, S. S., Kuy, S., Henry, T. D. & Patel, A. N. Cardiovascular disease, drug therapy, and mortality in covid-19. New England Journal of Medicine 382, e102 (2020).

[37] Jin, J.-M. et al. Gender differences in patients with COVID-19: Focus on severity and mortality. Frontiers in Public Health 8, 152 (2020).

[38] Petrakis, D. et al. Obesity – a risk factor for increased COVID-19 prevalence, severity and lethality (review). Molecular medicine reports 22, 9–19 (2020).

[39] Lighter, J. et al. Obesity in patients younger than 60 years is a risk factor for covid-19 hospital admission. Clinical infectious diseases: an official publication of the Infectious Diseases Society of America (2020).

[40] Flegal, K. M., Carroll, M. D., Kit, B. K. & Ogden, C. L. Prevalence of Obesity and Trends in the Distribution of Body Mass Index Among US Adults, 1999-2010. JAMA 307, 491–497 (2012).

[41] Guo, W. et al. Diabetes is a risk factor for the progression and prognosis of COVID-19. Diabetes/Metabolism Research and Reviews n/a, e3319 (2020).

[42] Wang, B., Li, R., Lu, Z. & Huang, Y. Does comorbidity increase the risk of patients with COVID-19: evidence from meta-analysis. Aging 12, 6049–6057 (2020).

[43] Yan, Y. et al. Clinical characteristics and outcomes of patients with severe covid-19 with diabetes. BMJ Open Diabetes Research and Care 8 (2020).

[44] Cheng, Y. et al. Kidney disease is associated with inhospital death of patients with COVID-19. Kidney International 97, 829–838 (2020).

[45] Emami, A., Javanmardi, F., Pirbonyeh, N. & Akbari, A. Prevalence of underlying diseases in hospitalized patients with COVID-19: a systematic review and metaanalysis. Archives of academic emergency medicine 8, e35–e35 (2020).

[46] Henry, B. M. & Lippi, G. Chronic kidney disease is associated with severe coronavirus disease 2019 (covid-19) infection. International Urology and Nephrology 52, 1193–1194(2020).

[47] Bansal, M. Cardiovascular disease and covid-19. Diabetes & Metabolic Syndrome: Clinical Research & Reviews 14, 247–250 (2020).

[48] Guo, T. et al. Cardiovascular Implications of Fatal Outcomes of Patients With Coronavirus Disease 2019 (COVID-19). JAMA Cardiology (2020).

[49] Zheng, Y.-Y., Ma, Y.-T., Zhang, J.-Y. & Xie, X. COVID-19 and the cardiovascular system. Nature Reviews Cardiology 17, 259–260 (2020).

[50] Shahid, Z. et al. COVID-19 and older adults: What we know. Journal of the American Geriatrics Society 68, 926–929 (2020).

[51] Yancy, C. W. COVID-19 and African Americans. JAMA 323, 1891–1892 (2020).

[52] Dyer, O. Covid-19: Black people and other minorities are hardest hit in US. BMJ 369, m1483 (2020).

[53] Zhao, X. et al. Incidence, clinical characteristics and prognostic factor of patients with COVID-19: a systematic review and meta-analysis. *medRxiv* (2020). Preprint at https://www.medrxiv.org/content/10.1101/2020.03.17.20037572v1.

[54] Li, K. et al. The clinical and chest CT features associated with severe and critical COVID-19 pneumonia. Investigative radiology 55, 327–331 (2020).

[55] Asadi-Pooya, A. A. & Simani, L. Central nervous system manifestations of COVID-19: A systematic review. Journal of the Neurological Sciences 413, 116832 (2020).

[56] Kim, H.-S., Lee, S. & Kim, J. H. Real-world evidence versus randomized controlled trial: Clinical research based on electronic medical records. Journal of Korean medical science 33, e213–e213 (2018).

[57] Bachtiger, P., Peters, N. S. & Walsh, S. L. Machine learning for COVID-19—asking the right questions. The Lancet Digital Health (2020).

[58] Watson Health, IBM Corporation. Ibm explorys network—unlock the power of big data beyond the walls of your organization. Tech. Rep., Route 100, Somers, NY 10589 (2016).

[59] Angelini, D. E., Radivoyevitch, T., McCrae, K. R. & Khorana, A. A. Bleeding incidence and risk factors among cancer patients treated with anticoagulation. American Journal of Hematology 94, 780–785 (2019).

[60] Kaelber, D. C., Foster, W., Gilder, J., Love, T. E. & Jain, A. K. Patient characteristics associated with venous thromboembolic events: a cohort study using pooled electronic health record data. Journal of the American Medical Informatics Association 19, 965–972 (2012).

[61] Pfefferle, K. J., Shemory, S. T., Dilisio, M. F., Fening, S. D. & Gradisar, I. M. Risk factors for manipulation after total knee arthroplasty: A pooled electronic health record database study. The Journal of Arthroplasty 29, 2036–2038 (2014).

[62] Raket, L. L. et al. Dynamic ElecTronic hEalth reCord deTection (DETECT) of individuals at risk of a first episode of psychosis: a case-control development and validation study. The Lancet Digital Health 2, e229–e239 (2020).

[63] Ravizza, S. et al. Predicting the early risk of chronic kidney disease in patients with diabetes using real-world data. Nature Medicine 25, 57–59 (2019).

[64] LOINC. SARS Coronavirus 2 – LOINC (2020). Accessed on 2020-04-20 https://loinc.org/sars-coronavirus-2/.

[65] Wang, W., Tang, J. & Wei, F. Updated understanding of the outbreak of 2019 novel coronavirus (2019-ncov) in wuhan, china. Journal of Medical Virology 92, 441–447 (2020).

[66] Dormann, C. F. et al. Collinearity: a review of methods to deal with it and a simulation study evaluating their performance. Ecography 36, 27–46 (2013).

[67] Olson, R. S., La Cava, W., Mustahsan, Z., Varik, A. & Moore, J. H. Data-driven advice for applying machine learning to bioinformatics problems. In Biocomputing 2018, 192–203 (2018).

[68] Van Calster, B., McLernon, D. J., Van Smeden, M., Wynants, L. & Steyerberg, E. W. Calibration: the achilles heel of predictive analytics. BMC Medicine 17, 230 (2019).

